# Automated Gland MRI Radiomics for Predicting Time to Pathologic Progression during Prostate Cancer Active Surveillance

**DOI:** 10.64898/2026.11.17.26368266

**Authors:** Hongye Zeng, Shreeram Athreya, Dingyuan Dai, Mara Pleasure, Vedrana Ivezić, Steve Raman, Leonard Marks, William Speier, Corey Arnold

## Abstract

**Background:** Repeated biopsies burden patients during prostate cancer active surveillance (AS). Existing MRI risk stratification tools avoiding biopsy remain limited by the difficulty of automated lesion segmentation.

**Purpose:** To develop a model integrating gland MRI radiomics and clinicopathologic variables for predicting time to pathologic progression during prostate cancer AS.

**Materials and Methods:** This retrospective single-institution study included men with grade group 1 or favorable grade group 2 prostate cancer undergoing AS with fusion-targeted and systematic biopsy (2010–2025). Gland segmentation used nnU-Net; gland radiomic features were selected by LASSO-penalized survival regression to train an XGBoost accelerated failure time model. Comparator models were assessed by C-index, time-dependent area under the receiver operating characteristic curve (AUC), Kaplan–Meier, and decision curve analysis.

**Results:** Of 522 men (median age, 64 years [IQR, 59–68]), 410 were assigned by site to development and 112 to internal testing. Gland segmentation Dice was 0.89 ± 0.04. The C-index was higher for the gland radiomics model (0.77) than for clinicopathologic (0.68) and lesion radiomics (0.68) models (adjusted P ≤ .017), with the highest time-dependent AUC at 36 months (0.87 [95% CI: 0.78, 0.94]). Progression-free survival was lower in high-risk than low-risk groups (36 months, 63.9% vs 100%; 48 months, 55.4% vs 93.8%; log-rank P < .001). At a 0.25 risk threshold, 49.1% and 31.3% were low risk at 36 and 48 months (negative predictive value, 1.00; positive predictive value, 0.36).

**Conclusion:** An automated gland radiomics model predicted time to pathologic progression during AS better than clinicopathologic and lesion-based models.

## Introduction

Prostate cancer is one of the most frequently diagnosed malignancies in men worldwide and a leading cause of cancer mortality in the United States (1). Approximately two-thirds of patients are eligible for monitoring, and active surveillance (AS) is now standard for low- and favorable intermediate-risk disease, deferring the morbidity of overtreatment while maintaining oncologic safety (2,3). However, AS protocols rely heavily on repeated prostate biopsies, which cause discomfort and carry risks of severe infection, pain, and bleeding (4,5). Notably, 66%–90% of repeat biopsies do not demonstrate histopathologic prostate cancer progression and may therefore be considered unnecessary (6,7). Reducing biopsy frequency could lower surveillance dropout and improve long-term adherence to AS.

Several personalized risk calculators incorporating clinicopathologic variables, including prostate-specific antigen (PSA), PSA density, and biopsy outcomes, have been developed to guide AS management (8–12). Subsequent integration of reader-dependent MRI assessments, including Prostate Imaging Reporting and Data System (PI-RADS) scores and Prostate Cancer Radiological Estimation of Change in Sequential Evaluation (PRECISE) ratings, has yielded incremental improvements in risk-stratification accuracy (13–15). More recently, radiomic and deep learning approaches have been applied to AS cohorts (16–22), leveraging quantitative imaging features to predict disease progression and yielding results comparable to or exceeding conventional models. These radiomics and deep learning approaches share several fundamental limitations. Lesion-based radiomic approaches are constrained by operator-dependent manual delineation, and fully automated alternatives remain unreliable due to the inherent difficulty of delineating small or inapparent lesions in low-risk prostate cancer on MRI (23,24). Beyond segmentation, most existing models predominantly address progression prediction as a binary classification problem at fixed time points, precluding estimation of time to progression and limiting their utility for individualized surveillance planning (16–20). Moreover, evidence is largely derived from small cohorts, raising concerns about generalizability.

To address these limitations, this study aimed to develop a survival prediction model for pathologic progression during prostate cancer AS by integrating gland MRI radiomics with clinicopathologic variables, without relying on lesion segmentation. Using data from eight imaging sites within a single academic medical system, we evaluated the model’s predictive performance and clinical utility in an independent internal test set.

## Materials and Methods

### Patient Population

This single-institution study across eight imaging sites was a retrospective analysis of prospectively acquired data, approved by our Institutional Review Board. We included all men prospectively and consecutively enrolled in an institutional AS program between 2010 and 2025 with prostate cancer confirmed by MR-US fusion–targeted and systematic biopsy (Artemis Inc, Eigen, USA). Biopsies were performed with coordinate memory to enable serial spatially localized sampling. All imaging data were de-identified prior to analysis by removal of protected health information (including patient name and medical record number) from DICOM metadata. Specific inclusion criteria were: (a) histologically confirmed grade group (GG) 1, or GG 2 prostate cancer with favorable histopathologic features on biopsy (Gleason 3+4 with limited pattern 4 component); and (b) at least two AS time points with available biopsy results. Exclusion criteria were: (a) prior definitive treatment, (b) incomplete biparametric MRI (bpMRI) data, and (c) failed gland segmentation. Pathologic progression was defined as upgrading to a higher GG on serial memory-based surveillance biopsy. Patients without progression were censored at the date of their last available biopsy. All eligible patients were split by imaging site into a development set and an internal test set. The patient selection process is summarized in Figure 1.

**Figure 1.**
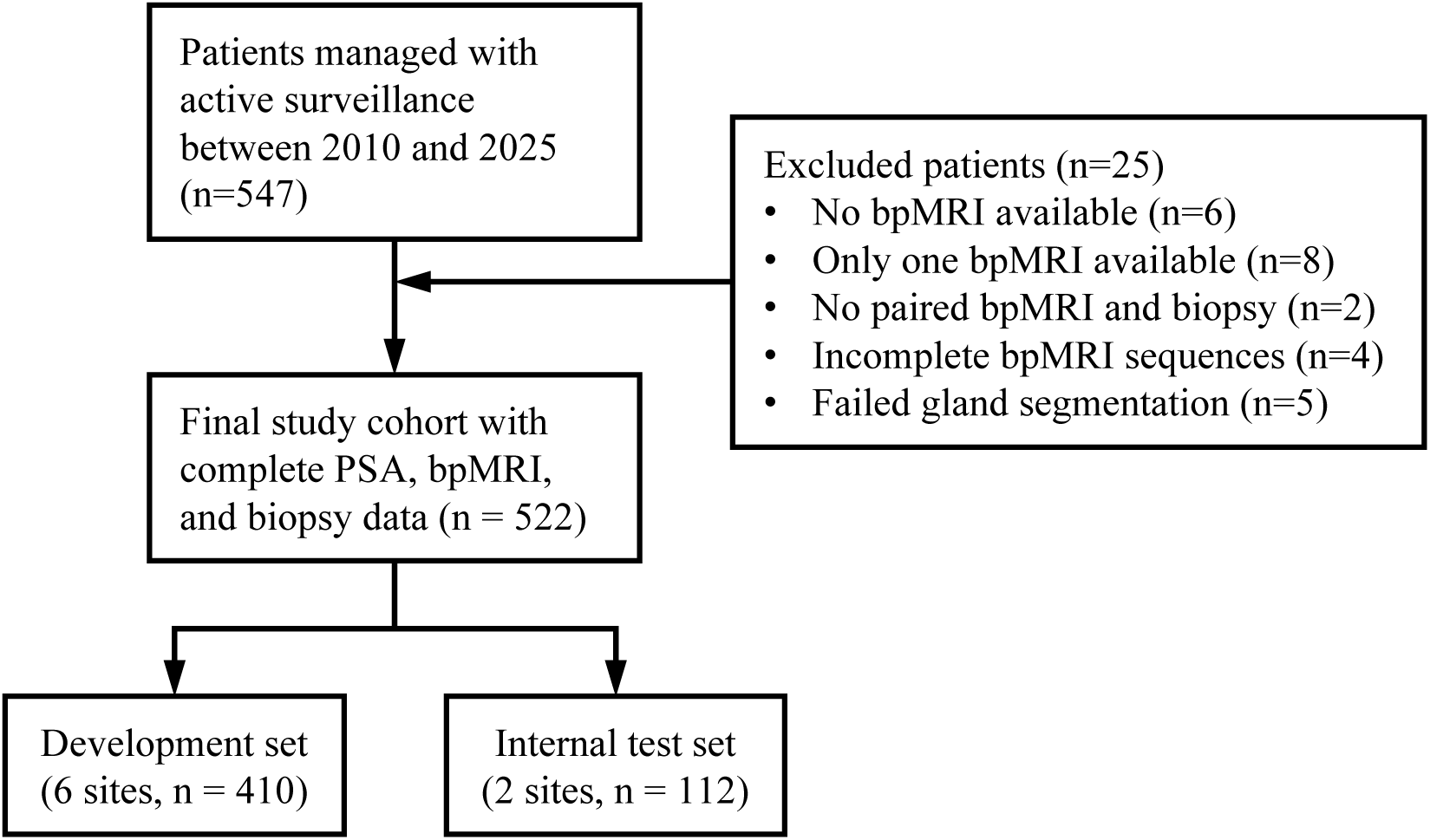
Patient flowchart for the retrospective cohort of men undergoing active surveillance for prostate cancer.

**Figure 2.**
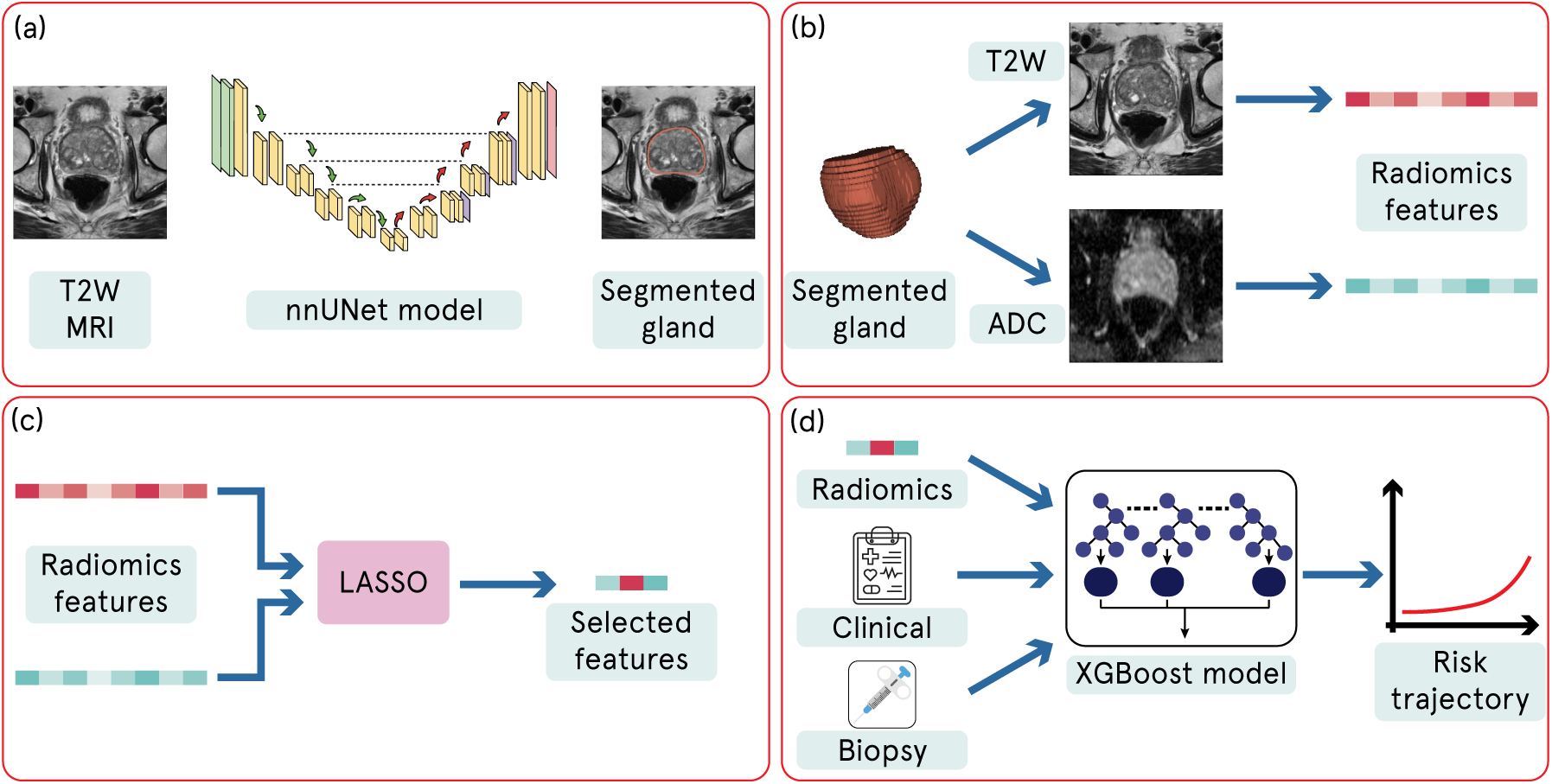
Overview of the multimodal risk stratification pipeline. Gland prostate segmentation was performed automatically using nnU-Net on T2-weighted images. Radiomic features were extracted from the segmented gland, reduced by Least Absolute Shrinkage and Selection Operator (LASSO) with stability selection. An XGBoost accelerated failure time model was then trained to predict time to pathologic progression.

### MRI Acquisition

All bpMRI examinations were performed primarily on 3-T scanners (or 1.5 T scanners in patients with orthopedic hardware) (Siemens Healthineers, Erlangen, Germany) with pelvic phased array coils at eight imaging sites within a single academic medical system. The bpMRI protocol included T2-weighted (T2W) and diffusion-weighted sequences with previously published protocols optimized for each scanner, exceeding the Prostate Imaging Reporting and Data System version 2.1 (PI-RADS v2.1) standards across the study period, reflecting the 15-year retrospective timeframe. All scans were centrally monitored and read by a group of prostate specialized academic abdominal radiologists. Diffusion-weighted imaging was acquired with b-values including 0 and 1000 s/mm², with calculated b-values exceeding 1400 s/mm²; apparent diffusion coefficient (ADC) maps were computed from acquired b-values. Full acquisition parameters are provided in Appendix S1 (Supplementary Methods 1).

### Segmentation model development

Prostate gland segmentation was performed automatically using nnU-Net (25) trained on the publicly available PI-CAI dataset (1,471 patients) (24), which was independent of the study cohort. The 3D full-resolution nnU-Net configuration (v2) was applied to the T2W series, and ADC maps were resampled to the T2W geometry so that a single gland mask was applied to both sequences. Lesion segmentation was also generated automatically using a model trained on the same PI-CAI dataset, with both the T2W and ADC series supplied as input channels. Both pipelines were therefore derived from identical training data. In an external subset of 100 patients sampled across all imaging sites, gland segmentation was validated against manual prostate contours drawn by two prostate-specialized academic abdominal radiologists; lesion detection was evaluated against patient-level reference labels. Both validations are detailed in Appendix S1.

### Radiomic Feature Extraction and Selection

A total of 200 radiomics features were extracted from the gland ROI on each image sequence (T2W and ADC) using PyRadiomics (version 3.1.0; https://github.com/AIM-Harvard/pyradiomics). To refine the feature set for the survival model, LASSO-penalized Cox regression with fold-wise stability selection was applied in the development set. The detailed process of feature extraction and selection is described in Appendix S1 (Supplementary Methods 3). Ultimately, the three radiomics features meeting the stability threshold were used to train the survival model (see Table S3).

### Survival Model Development and Validation

An XGBoost (eXtreme Gradient Boosting) survival model was trained using an accelerated failure time formulation (26). Model input comprised the selected gland radiomics features together with clinicopathologic variables (age, PSA, PSA density, GG, positive core percentage, and maximum tumor core). Hyperparameters were selected using five-fold cross-validation performed only on the development set, with patients stratified by event status; further details are provided in Appendix S1 (Supplementary Methods 4). The final model was then validated on the held-out internal test set. Two comparator models were trained under identical conditions using the same XGBoost survival model architecture: a clinicopathologic model (clinicopathologic variables only) and a lesion radiomics model (clinicopathologic variables plus lesion-level radiomics features).

### Statistical Analysis

Model discrimination in the internal test set was assessed using the concordance index (C-index) as the primary endpoint. Time-dependent AUCs at 24, 36, 48, and 60 months were reported as secondary endpoints. C-index and time-dependent AUC were reported with 95% confidence intervals (CI) estimated by bootstrap resampling (1,000 iterations). Differences in C-index and time-dependent AUC between models were assessed by paired bootstrap on the same resamples, with two-sided P values derived from the bootstrap distribution of the difference. To account for multiple comparisons across models and time points, P values were adjusted using the Benjamini-Hochberg procedure to control the false discovery rate at .05.

Clinical utility was assessed using decision curve analysis, comparing the model’s net benefit with the biopsy-all and biopsy-none strategies. For Kaplan–Meier analysis, patients were dichotomized into high- and low-risk groups at the median predicted risk score of the development set, and this fixed threshold was applied unchanged to the internal test set. Kaplan–Meier curves were constructed to visualize time to progression by risk group, with between-group differences assessed using the log-rank test. Calibration was evaluated using calibration curves. All statistical analyses were performed in Python (v3.9), using the dcurves and lifelines packages for decision curve analysis and survival analysis, respectively. Sample size was based on all eligible patients identified within the study period; no formal power analysis was performed.

## Results

### Characteristics of the Study Cohort

A total of 547 patients were eligible, of whom 25 were excluded, resulting in a final study cohort of 522 patients (Figure 1). Table 1 presents the baseline characteristics of the internal development (n = 410) and internal testing (n = 112) cohorts. Median age (63 vs 65 years), PSA (5.7 vs 5.8 ng/mL), and PSA density (0.11 ng/mL² in both) were similar between the development and testing cohorts. Biopsy findings at the baseline visit were likewise similar, with median positive core percentages of 11.8% (IQR, 5.9–23.4) and 12.5% (IQR, 6.3–23.9) and median maximum tumor percentages of 20% (IQR, 5–40) in both cohorts. Grade group 1 disease was present in 75.6% (310 of 410) and 76.8% (86 of 112) of patients in the development and testing cohorts, respectively. At the end of follow-up, 23.9% (98 of 410) and 24.1% (27 of 112) of patients had experienced pathologic progression, with median times to event of 35.2 months (IQR, 15.1–57.3 months) and 35.8 months (IQR, 21.8–62.4 months), respectively.

**Table 1.**
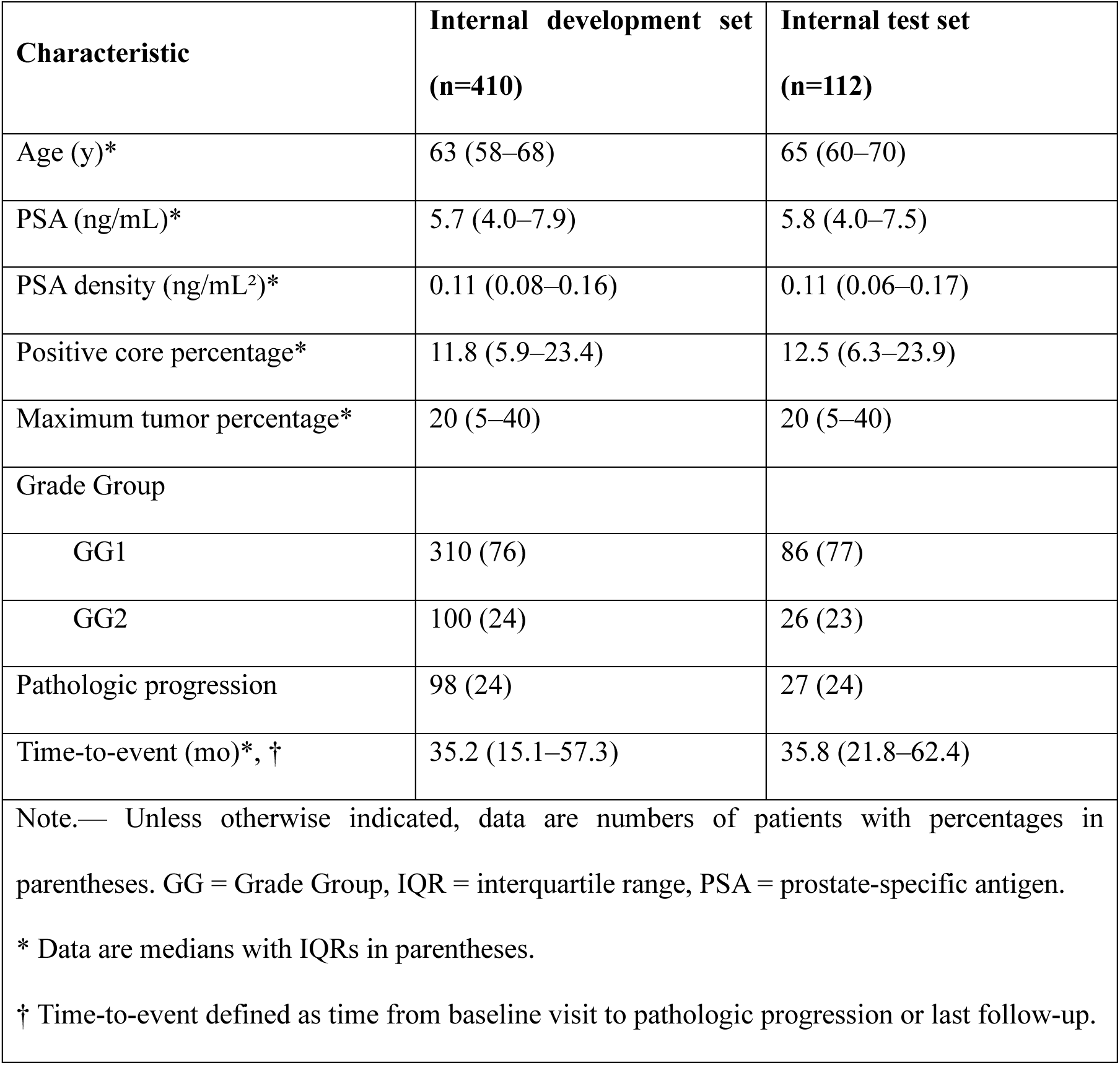
Characteristics of the Study Cohort.

| **Characteristic** | **Internal development set (n=410)** | **Internal test set**<br>**(n=112)** |
|---|---|---|
| Age (y)* | 63 (58–68) | 65 (60–70) |
| PSA (ng/mL)* | 5.7 (4.0–7.9) | 5.8 (4.0–7.5) |
| PSA density (ng/mL<sup>2</sup>)* | 0.11 (0.08–0.16) | 0.11 (0.06–0.17) |
| Positive core percentage* | 11.8 (5.9–23.4) | 12.5 (6.3–23.9) |
| Maximum tumor percentage* | 20 (5–40) | 20 (5–40) |
| Grade Group |  |  |
| GG1 | 310 (76) | 86 (77) |
| GG2 | 100 (24) | 26 (23) |
| Pathologic progression | 98 (24) | 27 (24) |
| Time-to-event (mo)*, † | 35.2 (15.1–57.3) | 35.8 (21.8–62.4) |

Note.— Unless otherwise indicated, data are numbers of patients with percentages in parentheses. GG = Grade Group, IQR = interquartile range, PSA = prostate-specific antigen.
\* Data are medians with IQRs in parentheses.
† Time-to-event defined as time from baseline visit to pathologic progression or last follow-up.

### Segmentation Performance

Segmentation performance is summarized in Tables S1 and S2. In internal five-fold cross-validation on the PI-CAI dataset, automated gland segmentation reached a Dice similarity coefficient of 0.96 (standard deviation (SD) < 0.01) and an intersection over union (IoU) of 0.92 (SD < 0.01). In the external subset of 100 patients, agreement remained high, with a Dice of 0.89 ± 0.04 and an IoU of 0.81 ± 0.06 (Table S1). The model was then applied to the entire study cohort. Segmentation failed in 5 patients, in whom the output mask was empty. All 5 had axial T2W images acquired at a large slice thickness of 3 mm or 5 mm. These 5 patients were excluded from the development set. Representative failure cases are shown in Figure S1.

In the same five-fold cross-validation on the PI-CAI dataset, lesion segmentation performed markedly worse, with a Dice of 0.208 ± 0.018 and an IoU of 0.159 ± 0.014. As described in Appendix S1, we therefore validated the model in the same subset using whether a lesion was successfully detected as the criterion. Among the 41 patients with a reference-positive lesion, the model detected a lesion in only 15 (sensitivity 0.37). Full detection metrics are reported in Table S2.

### Survival Model Performance Comparison

Survival model performance is summarized in Table 2. The gland radiomics model achieved the highest discrimination (C-index, 0.77 [95% CI: 0.70, 0.85]), outperforming both the clinicopathologic and lesion radiomics models (C-index, 0.68 for both; adjusted P = .016 and .017, respectively). Time-dependent AUC was consistently higher for the gland radiomics model at every evaluated time point (range, 0.83 to 0.87), with the largest value at 36 months (0.87 [95% CI: 0.78, 0.94]). After adjustment for multiple comparisons (Table S4), differences against the clinicopathologic model remained significant at all time points (P ≤ .020), and differences against the lesion radiomics model remained significant at 24, 36, and 60 months (P ≤ .038) but were borderline at 48 months (P = .050). The clinicopathologic and lesion radiomics models performed similarly throughout follow-up (AUC range, 0.70 to 0.76). Brier scores were consistently lower for the gland radiomics model across all evaluated time points, indicating lower overall prediction error than the clinicopathologic and lesion radiomics models (Table S5). Calibration curves comparing predicted and observed progression-free probabilities across the evaluated time points are shown in Figure S2.

**Table 2.** Performance of the Whole-Gland Radiomics Model Compared with Clinical and Lesion Radiomics Models.

| Metric | Clinicopathologic | Lesion radiomics | Gland radiomics | P (vs Clinical) | P (vs Lesion) |
|---|---|---|---|---|---|
| C-index | 0.68<br>(0.57, 0.79) | 0.68<br>(0.58, 0.79) | 0.77<br>(0.70, 0.85) | .016 | .017 |
| AUC@24m | 0.70<br>(0.54, 0.84) | 0.70<br>(0.56, 0.84) | 0.83<br>(0.72, 0.91) | <.001 | .010 |
| AUC@36m | 0.73<br>(0.59, 0.86) | 0.75<br>(0.63, 0.87) | 0.87<br>(0.78, 0.94) | <.001 | .007 |
| AUC@48m | 0.75<br>(0.60, 0.87) | 0.76<br>(0.61, 0.87) | 0.85<br>(0.73, 0.94) | .020 | .050 |
| AUC@60m | 0.74<br>(0.59, 0.86) | 0.76<br>(0.61, 0.87) | 0.85<br>(0.72, 0.93) | .020 | .038 |

Note.—Data in parentheses are 95% CIs. C-index and time-dependent AUC were estimated by bootstrap resampling (1,000 iterations). Two-sided P values were derived from the bootstrap distribution of the difference between the gland radiomics model and the indicated comparator model. P values were adjusted for multiple comparisons (n = 10) using the Benjamini-Hochberg procedure. Unadjusted P values and the full adjustment are reported in Table S4.

### Clinical Utility Analysis

Decision curve analysis demonstrated the net benefit of the gland radiomics model over biopsy-all and biopsy-none strategies at 36 and 48 months (Figure 3). Within the threshold range of approximately 0.15–0.30, the gland radiomics model consistently showed higher net benefit than the clinicopathologic and lesion radiomics models. Kaplan–Meier analysis demonstrated significantly worse progression-free survival in the high-risk group compared with the low-risk group (log-rank P < .001; Figure 4). Progression-free survival estimates were 63.9% in the high-risk group versus 100% in the low-risk group at 36 months, and 55.4% versus 93.8% at 48 months. To translate these findings into a clinically actionable estimate, we quantified the proportion of patients who could potentially avoid repeat biopsy at model-defined risk thresholds within the range supported by the decision curve analysis (Table 3). At a threshold of 0.25, 49.1% and 31.3% of patients were classified as low risk at 36 and 48 months, respectively, with corresponding negative predictive values (NPVs) of 1.00 at both time points, indicating no progression within the respective time horizon among patients who would have avoided biopsy. At a threshold of 0.30, the proportion classified as low risk increased to 58.0% and 40.2%, respectively, with a slight reduction in NPV (0.95 and 0.96, corresponding to missed-progression rates of 4.6% and 4.0%).

**Figure 3.**
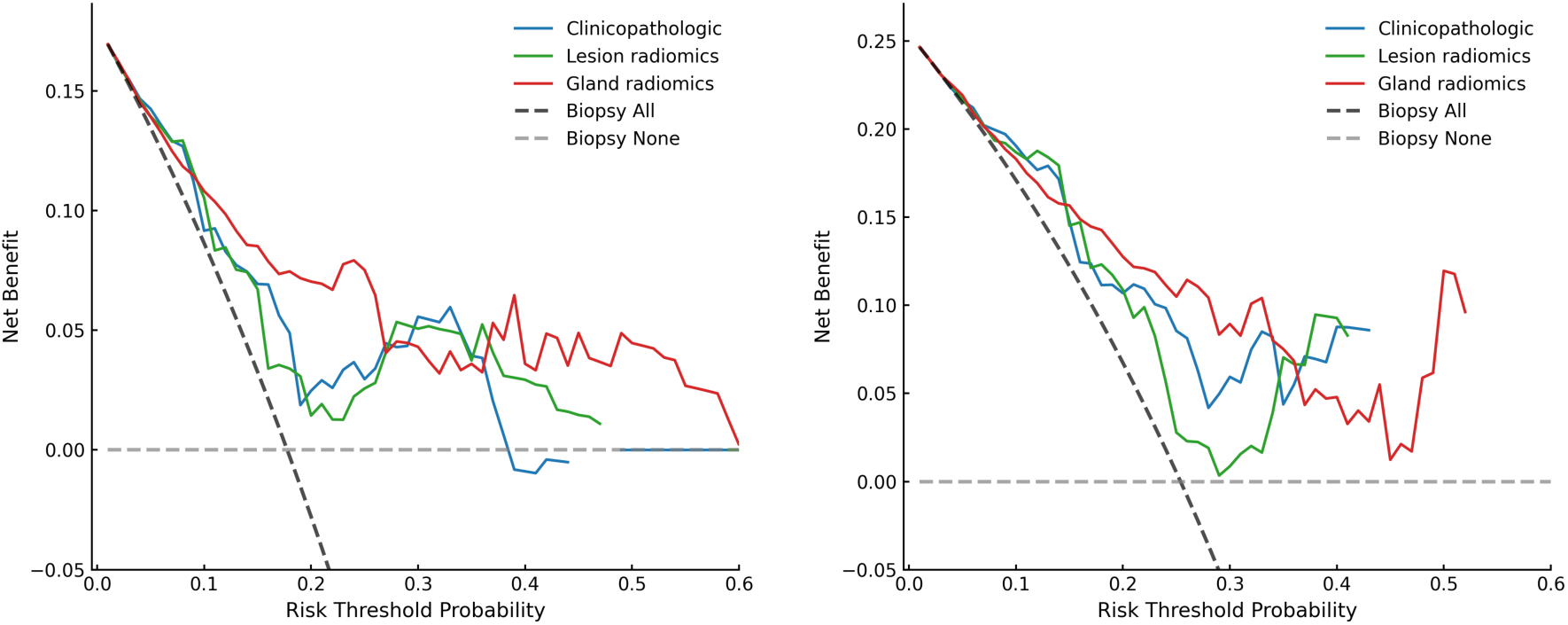
Decision curve analysis for the gland radiomics model and comparator models at (a) 36 and (b) 48 months. Net benefit is plotted as a function of threshold probability. Higher net benefit across clinically relevant threshold probabilities indicates greater clinical utility for personalizing biopsy frequency.

**Figure 4.**
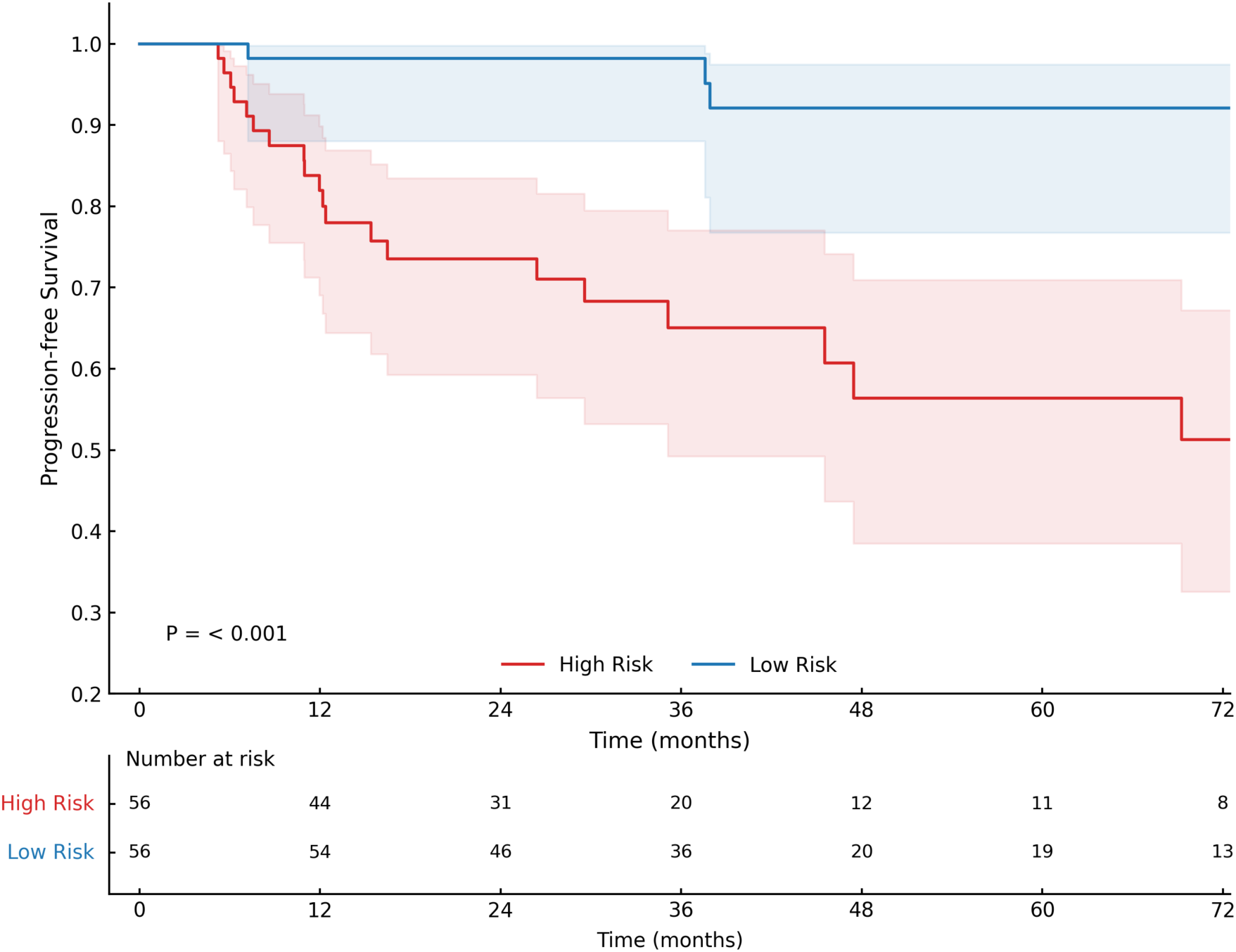
Kaplan–Meier curves of progression-free survival in the internal test set (n = 112), stratified into low- and high-risk groups by the gland radiomics model. The threshold was the median predicted risk score of the development set and was applied unchanged to the internal test set. Groups were compared with the log-rank test.

**Table 3.** Estimated Reduction in Biopsy Frequency at Model-Defined Risk Thresholds in the Internal Test Set.

| **Time** | **Risk threshold** | **% Low risk (biopsy avoidable)** | **NPV** | **PPV** |
|---|---|---|---|---|
| 36m | 0.25 | 49.1 (55/112) | 1.00 | 0.36 |
|  | 0.30 | 58.0 (65/112) | 0.95 | 0.37 |
| 48m | 0.25 | 31.3 (35/112) | 1.00 | 0.36 |
|  | 0.30 | 40.2 (45/112) | 0.96 | 0.41 |

Note.—Data in the low-risk column are percentages, with numerators and denominators in parentheses. Patients were classified as low risk when the model-predicted probability of pathologic progression at the indicated time point was below the corresponding risk threshold, in whom biopsy was considered avoidable. NPV = negative predictive value, PPV = positive predictive value.

### Model Interpretation

Feature importance analysis using Shapley additive explanations (SHAP) (27) values identified the T2W gland diameter as the most influential predictor, followed by age, positive core percentage, and grade group (Figure 5). Among clinicopathologic variables, older age and higher positive core percentage were associated with increased predicted risk, while a higher grade group was likewise associated with elevated risk. Among imaging features, T2W gland diameter showed a non-monotonic relationship with predicted risk, with only markedly enlarged glands associated with lower predicted risk; greater ADC texture complexity was associated with increased predicted risk. A representative case illustrating the clinical utility of the gland radiomics model is presented in Figure 6. In this patient, the gland radiomics model predicted a risk below the 0.25 threshold through 48 months, whereas the clinicopathologic and lesion radiomics models exceeded this threshold by 48 months; the patient remained progression-free until 99.9 months, beyond the evaluated window.

**Figure 5.**
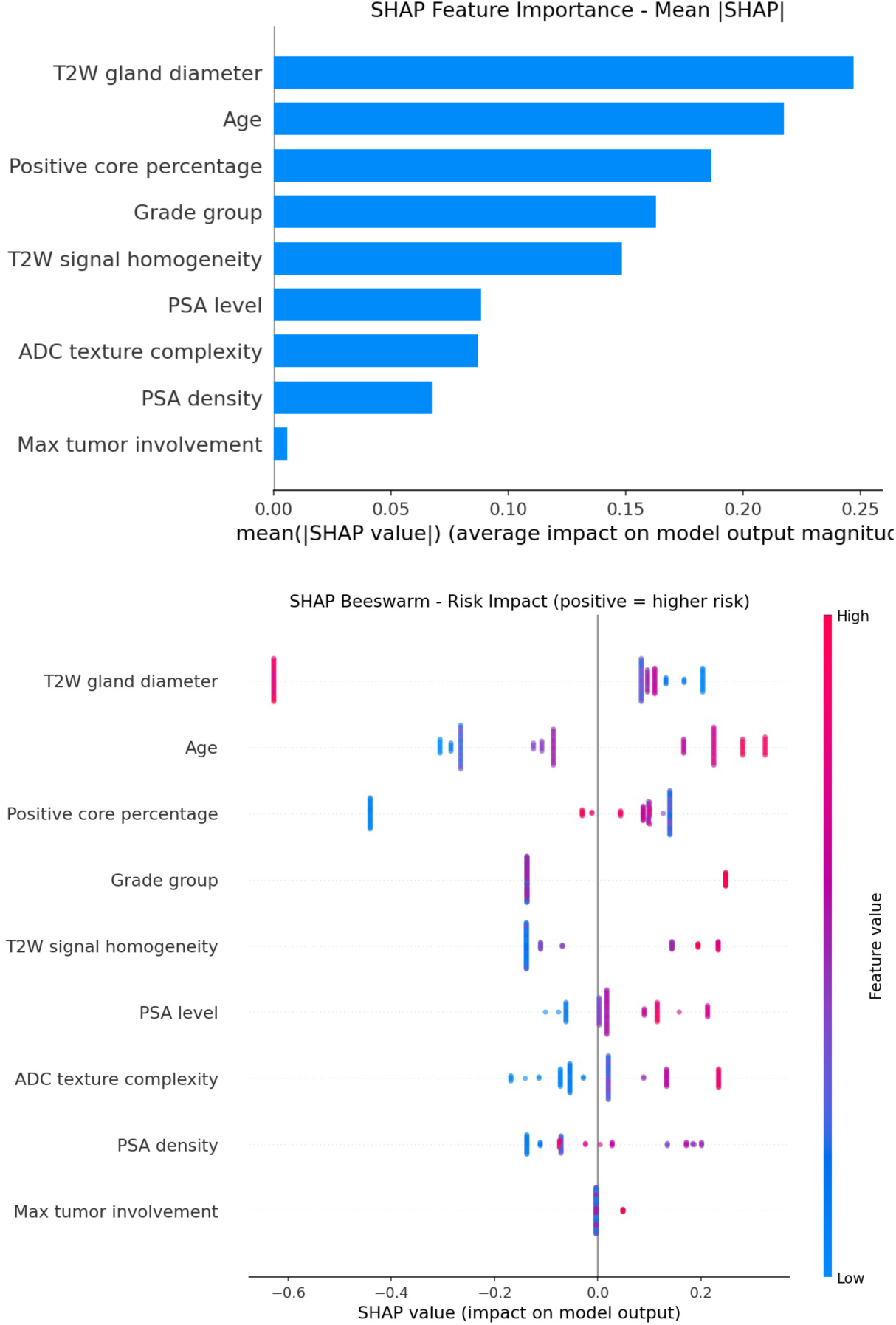
SHAP (SHapley Additive Explanations) analysis of the gland radiomics model. (a) Bar plot showing mean absolute SHAP values for each feature, ranked by overall contribution to model output. (b) Beeswarm plot illustrating the direction and magnitude of each feature’s impact on predicted risk, with color indicating feature value.

**Figure 6.**
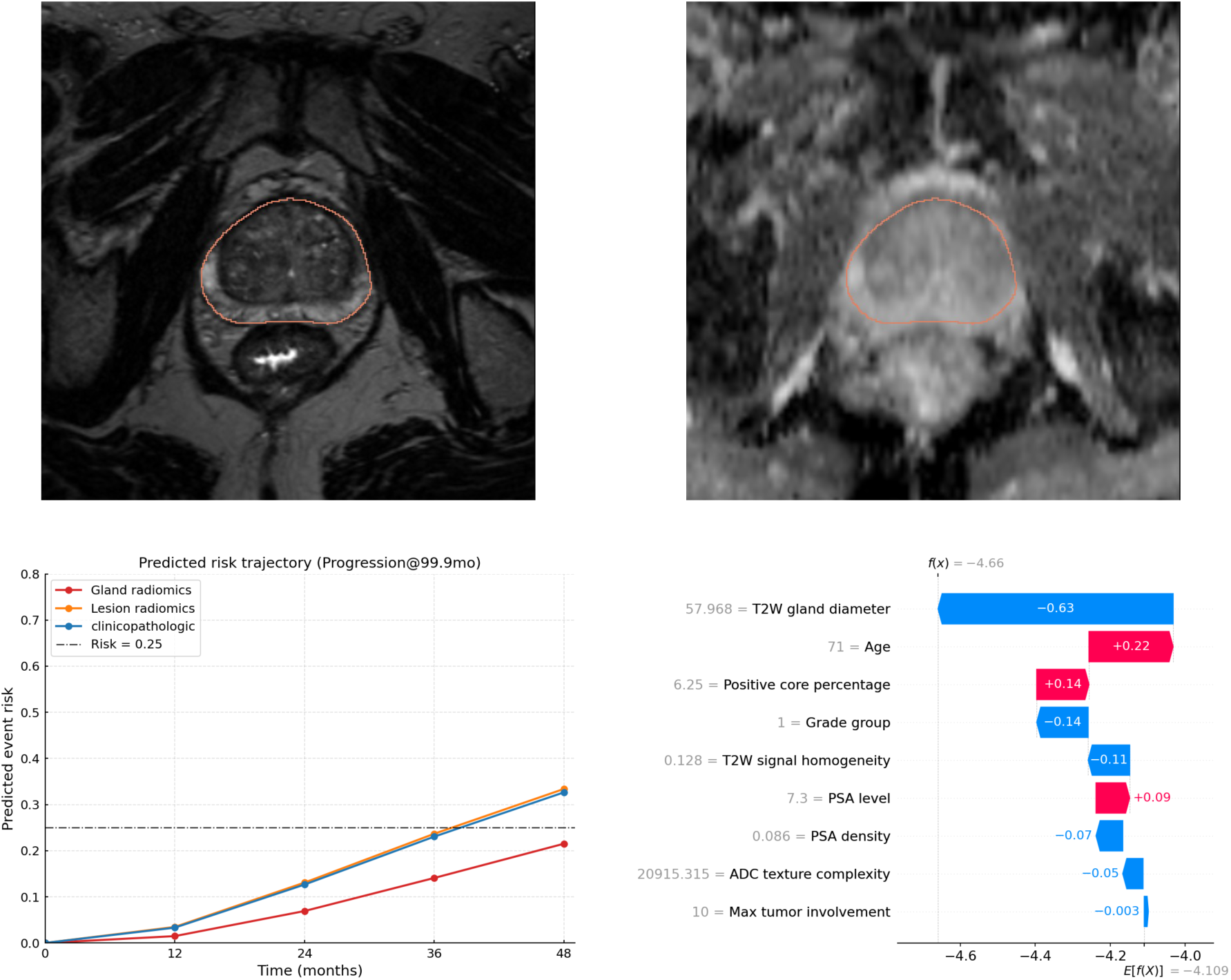
Illustrative case showing gland radiomics model behavior in a patient with pathologic progression at 99.9 months. This single case is provided for illustration only and is not evidence of model performance. (a, b) T2-weighted and axial ADC images demonstrate automated whole-gland segmentation (red contour). (c) Predicted risk trajectories over the follow-up period for the three comparator models; the dashed horizontal line indicates the 0.25 risk threshold. (d) SHAP waterfall plot showing the contribution of each feature to the gland radiomics model’s predicted risk for this patient, with feature values shown in their original units.

## Discussion

AS for prostate cancer requires accurate identification of patients at risk of pathologic progression to avoid both unnecessary biopsies and delayed intervention. In this retrospective study of 522 men undergoing AS, we developed a fully automated multimodal model that integrates gland MRI radiomics and clinicopathologic variables to predict the time to pathologic progression. In the internal test set, the model achieved a C-index of 0.77 (95% CI: 0.70, 0.85) and a time-dependent AUC of 0.87 (95% CI: 0.78, 0.94) at 36 months, outperforming both the clinicopathologic and lesion radiomics models (adjusted P ≤ .017). At a risk threshold of 0.25, the model classified 49.1% of patients as low risk at 36 months, with no pathologic progression observed within 36 months in this subgroup, suggesting that model-guided stratification may allow surveillance biopsies to be deferred in some patients.

Although time-to-event formulations are well established in clinical and reader-based AS risk models, prior MRI-based models have framed progression as a binary classification problem, whether using radiomic features or deep learning. Most predict whether progression has occurred within the current surveillance interval (16–20,22), and the one study that extended the horizon dichotomized patients into fast and slow progressors around a 12-month cutoff (21). These models were also developed in small cohorts, comprising fewer than 75 patients in radiomic studies dependent on manual lesion delineation (16–18,22), fewer than 210 patients in deep learning studies (19,20), and only 16 test patients in the study reporting the longest horizon, in which the AUC was 0.84 (21). By formulating progression as a time-to-event outcome in 522 patients with serial biopsy data, our present model estimated individualized progression-free probabilities across 24-to 60-month horizons in 112 held-out patients (time-dependent AUC, 0.87 at 36 months), extending MRI-based progression prediction from a single-interval label to a continuous estimate of progression timing.

A further limitation of prior MRI-based models is their dependence on lesion segmentation. Lesion delineation is intrinsically difficult, with state-of-the-art automated methods reporting Dice similarity coefficients of approximately 0.4–0.5 (23,24,28); in the present implementation, lesion segmentation reached a Dice of only 0.21 in cross-validation and detected a lesion in just 15 of 41 reference-positive patients when applied to our cohort (sensitivity, 0.37). Restricting a model to lesion-level features, therefore, introduces selection bias by excluding AS patients in whom no lesion is visible on MRI, yet systematic biopsy nonetheless reveals a higher grade group. Gland segmentation, by contrast, is substantially more reproducible and amenable to full automation across institutions (29,30). Developed on the European multicenter PI-CAI dataset, the gland model transferred to the present institutional cohort with only a modest loss of accuracy relative to internal cross-validation (Dice, 0.96 vs 0.89), and produced an empty mask in only 5 patients, all acquired at 3-or 5-mm slice thickness.

The intended use is decision support at each surveillance visit, to inform whether the next scheduled biopsy can be deferred. At a risk threshold of 0.25, 49.1% of patients at 36 months fell into a group with no progression, whereas a threshold of 0.30 classified 58.0% as low risk, at the cost of missing 4.6% of events. Positive predictive values were modest (0.36); the model is therefore suited to identifying patients in whom surveillance biopsy might be deferred, not to confirming patients who require intervention. Kaplan–Meier and decision curve analyses support this separation and show greater net benefit for model-guided selection than for biopsying all or no patients over clinically relevant thresholds. However, no patient’s schedule was altered in this retrospective cohort, so both the threshold and the safety of any resulting deferral require prospective evaluation, and the widening confidence intervals beyond 48 months call for caution at later time points.

This study has several limitations. First, the prognostic model was evaluated only in a held-out subset of a single retrospective cohort; although the gland segmentation model was validated in an external dataset, the prognostic model was not, and external validation is needed before clinical use. Second, progression was defined as a grade group upgrade on combined MRI-targeted and systematic biopsy, which is subject to sampling error, so an apparent upgrade may reflect detection of pre-existing higher-grade disease rather than true biologic progression, and conversely, some progression events were likely missed. Third, progression was ascertained only at biopsy, so the true time of upgrade lies somewhere within the interval between biopsies, and treating the biopsy date as the event time imposes a degree of interval censoring that the present survival framework does not explicitly model. Fourth, biopsy timing was clinically determined rather than protocol-fixed, and patients with greater clinical suspicion were likely biopsied sooner, so ascertainment was probably informative with respect to risk and may bias estimated progression times. Inter- and intra-rater variability of the reference gland contours was not formally assessed, which may contribute residual reference-standard uncertainty.

In conclusion, a fully automated gland MRI radiomics model predicted the time to pathologic progression during prostate cancer AS more accurately than clinicopathologic or lesion-based models. Gland segmentation proved more robust across datasets than lesion segmentation, and its whole-gland target allowed the model to be applied irrespective of lesion visibility. External and prospective validation are needed before it can inform biopsy scheduling.

## Data Availability

The MRI and clinical data used in this study are not publicly available due to patient privacy and institutional restrictions. De-identified data may be available from the corresponding author upon reasonable request and with approval from the Institutional Review Board of the University of California, Los Angeles.

## Appendix S1: Supplementary Methods

### 1. MRI Protocol

Patients in the development set were imaged on 3.0T and 1.5T Siemens scanners (Skyra, TrioTim, Skyra_fit, Verio, MAGNETOM Vida, Prisma_fit, Avanto_fit). Patients in the hold-out test set were imaged on 3.0T and 1.5T Siemens scanners (Skyra, TrioTim, Prisma_fit, Avanto_fit, Avanto, MAGNETOM Vida, Verio, Skyra_fit). Two sequence images, T2-weighted (T2W) and apparent diffusion coefficient (ADC) maps derived from diffusion-weighted imaging, were evaluated from each patient’s first examination.

Parameters of T2W images in the development set were as follows: slice thickness, 1.50–5.00 mm; repetition time, mean 2466.4 ms, range 651.0–6750.0 ms; echo time, mean 188.0 ms, range 85.0–207.0 ms; flip angle, 90–160°; matrix, 256 × 256 to 736 × 512; in-plane resolution, 0.33 × 0.33 to 1.12 × 1.12 mm. Parameters of T2W images in the test set were as follows: slice thickness, 1.50–5.00 mm; repetition time, mean 2213.7 ms, range 700.0–4880.0 ms; echo time, mean 200.5 ms, range 85.0–207.0 ms; flip angle, 104–150°; matrix, 256 × 256 to 512 × 512; in-plane resolution, 0.33 × 0.33 to 1.19 × 1.19 mm.

Parameters of ADC maps in the development set were as follows: slice thickness, 3.00–4.60 mm; repetition time, mean 4700.9 ms, range 3200.0–8500.0 ms; echo time, mean 77.1 ms, range 52.0–99.0 ms; matrix, 96 × 96 to 256 × 256; in-plane resolution, 0.86 × 0.86 to 2.34 × 2.34 mm. Parameters of ADC maps in the test set were as follows: slice thickness, 3.00–3.60 mm; repetition time, mean 4542.6 ms, range 3200.0–5550.0 ms; echo time, mean 76.5 ms, range 51.0–99.0 ms; matrix, 110 × 108 to 160 × 132; in-plane resolution, 1.62 × 1.62 to 2.34 × 2.34 mm.

### 2. Segmentation model development and validation

All 1,471 patients in the PI-CAI dataset [1] were provided with expert segmentation masks for the whole prostate gland and for lesions. Both the gland and the lesion model were trained with nnU-Net using five-fold cross-validation. The gland model used the T2W image as its sole input channel, which was sufficient for delineating the entire prostate; the lesion model used both the T2W image and the ADC map as input channels, yielding better lesion segmentation. Preprocessing (resampling to 0.5 × 0.5 × 3.0 mm, and per-image z-score intensity normalization) was performed by the nnU-Net pipeline using its default configuration.

#### Gland segmentation validation

Internal performance was assessed by five-fold cross-validation on the PI-CAI dataset. For external validation, 100 patients were sampled across all imaging sites; each case was contoured by one of two prostate-specialized academic abdominal radiologists, whose annotations served as the reference standard. Ambiguous cases were adjudicated by a senior radiologist. Inter- and intra-rater variability was not formally assessed. Agreement was quantified by Dice and IoU in both validation settings. The model was then applied to our entire cohort. Results are reported in Table S1.

#### Lesion segmentation validation

Voxel-wise agreement was assessed using five-fold cross-validation on the PI-CAI dataset. Because voxel-wise lesion annotation is highly time-consuming, validation on our data was simplified to a lesion detection task within the same cohort of 100 patients: PI-RADS > 3 was used as the positive reference label (41 positive / 59 negative), and any model-segmented lesion was treated as a positive prediction. Detection performance was summarized at the patient level by sensitivity, specificity, accuracy, and F1 score. Results are reported in Table S2.

### 3. Radiomics extraction and selection

#### Region of interest

Whole-gland prostate masks were generated on T2W images using a 3D full-resolution nnU-Net (v2) segmentation model. T2W images, apparent diffusion coefficient (ADC) maps, and the corresponding gland mask were resampled to a common voxel spacing of 0.5 × 0.5 × 3.0 mm during the segmentation step. Because the three volumes were therefore already voxel-wise aligned at this spacing, no additional resampling was performed during the feature-extraction step.

#### MRI normalization

Because T2W signal intensity is a relative, non-quantitative unit that is not comparable across scanners/vendors, T2W voxel intensities were z-score normalized before gray-level discretization (normalize=True, normalizeScale=100, voxelArrayShift=300), and a fixed bin width of 5 was used, following the parameter set recommended in the official PyRadiomics. ADC maps are quantitative (mm^2^/s) and were left unnormalized per PyRadiomics guidance for quantitative images; default extraction settings were used (bin width = 25, no resampling).

#### Feature extraction

Radiomics features were extracted independently from the T2W images and ADC maps within the whole-gland mask using PyRadiomics (version 3.1.0) in Python 3. Features were computed only from the original image; no wavelet, Laplacian-of-Gaussian, square, or other derived-image filters were applied. For each sequence, seven feature classes were extracted: shape (3D, n=14), first-order statistics (n=18), gray-level co-occurrence matrix (GLCM, n=24), gray-level run-length matrix (GLRLM, n=16), gray-level size-zone matrix (GLSZM, n=16), gray-level dependence matrix (GLDM, n=14), and neighboring gray-tone difference matrix (NGTDM, n=5), for 107 features per sequence. Because the T2W images and ADC maps of a given patient share the same whole-gland mask, the 14 shape features are identical across the two sequences; the ADC shape features were therefore removed as duplicates, leaving 107 T2W features and 93 ADC features, i.e., 200 features per patient in total.

#### Feature selection

Features were selected in the development cohort using a LASSO-regularized Cox proportional-hazards model (CoxnetSurvivalAnalysis, scikit-survival). The development set was split into 5 patient-level folds, stratified by event status. The regularization strength (alpha) yielding the highest mean cross-validated C-index was selected. At this alpha, the model was refit within each fold, and the proportion of folds in which each feature received a non-zero coefficient was recorded as a stability measure. Features with a non-zero coefficient in at least 3 of the 5 folds were retained as the final feature set. The final selected features and their mean coefficients are listed in Table S3.

### 4. XGBoost hyperparameter

Hyperparameters for the XGBoost AFT model (max_depth, min_child_weight, and colsample_bytree) were jointly tuned via grid search in the development cohort, using the same patient-level, event-stratified 5-fold cross-validation scheme described above. The search space was max_depth ∈ {1, 2, 3}, min_child_weight ∈ {10, 20, 30}, and colsample_bytree ∈ {0.6, 0.8, 1.0}, yielding 27 candidate combinations. The AFT loss distribution was fixed to normal, and other hyperparameters were fixed at commonly used values. The combination yielding the highest mean cross-validated C-index in the development cohort was selected for the final model: XGB_PARAMS = { ’objective’: ’survival:aft’, ’eval_metric’: ’aft-nloglik’, ’aft_loss_distribution’: ’normal’, ’aft_loss_distribution_scale’: 1.0, ’max_depth’: 1, ’eta’: 0.05, ’min_child_weight’: 10, ’subsample’: 0.8, ’colsample_bytree’: 0.8, ’tree_method’: ’hist’}

## Appendix S2: Supplementary Results

### Gland segmentation performance

Dice and IoU in five-fold cross-validation on the PI-CAI dataset and in the external validation set of 100 patients are summarized in Table S1. When the model was applied to our entire cohort, segmentation failed in 5 patients, all of whose axial T2-weighted (T2W) images had a slice thickness of 3 mm or 5 mm. Representative failure cases are shown in Figure S1, including inadequate T2W field-of-view coverage that excluded the prostate and distortion of the posterior gland margin by an adjacent colonic diverticulum.

### Lesion segmentation and detection performance

Voxel-wise agreement for lesion segmentation was low in five-fold cross-validation (Dice = 0.208 ± 0.018; IoU = 0.159 ± 0.014). In the simplified lesion detection task in 100 patients, the confusion matrix is shown in Table S2. At the patient level, the model detected lesions with a sensitivity of 0.37, a specificity of 0.86, an overall accuracy of 0.66 and an F1 score of 0.47.

## Appendix S3: Supplementary Figures

**Figure S1.**
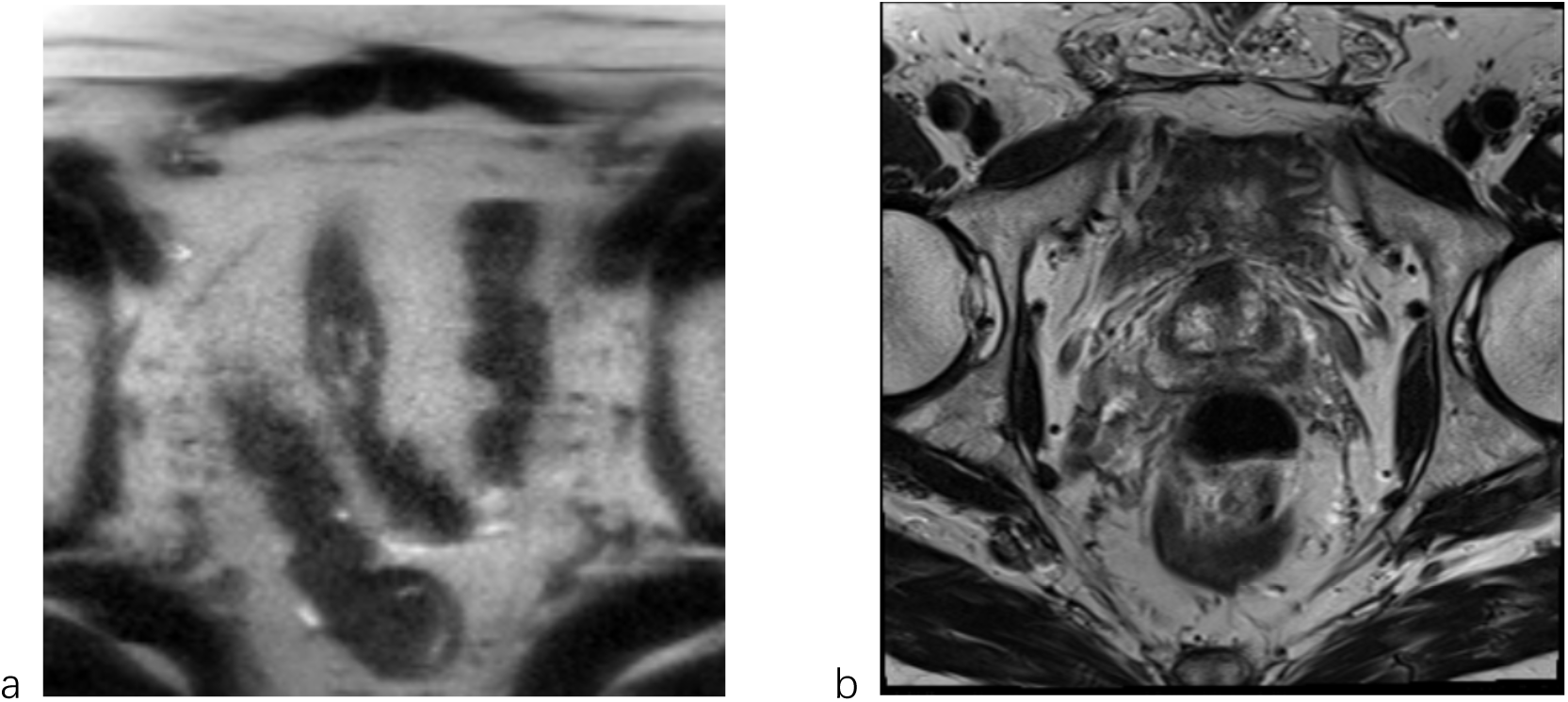
Representative segmentation failure cases. (a) Failure due to inadequate field-of-view coverage in the T2W sequence, excluding the prostate from the imaged volume. (b) Segmentation error due to an adjacent colonic diverticulum distorting the posterior prostate margin.

**Figure S2.**
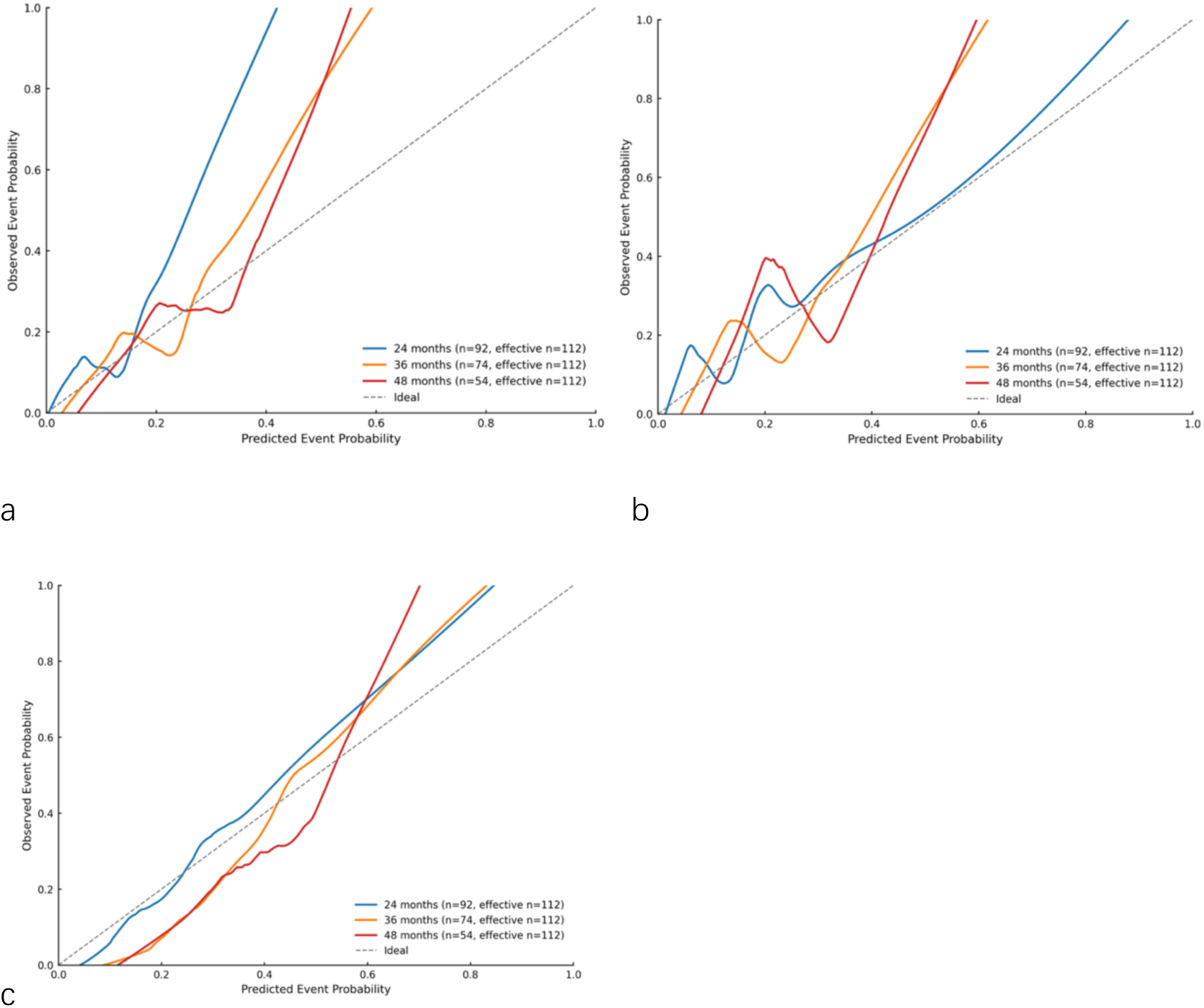
Calibration curves of the three models. (a) Clinicopathologic; (b) lesion radiomics; (c) gland radiomics.

## Appendix S4: Supplementary Tables

**Table S1.** Gland segmentation performance.

| **Validation** | **Dice (mean ± SD)** | **IoU (mean ± SD)** |
|---|---|---|
| Five-fold cross-validation (PI-CAI) | 0.959 ± 0.002 | 0.923 ± 0.002 |
| Our sampled data (n=100) | 0.891 ± 0.040 | 0.805 ± 0.061 |

**Table S2.** Lesion detection confusion matrix (n = 100)

|  | **Reference positive (PI-RADS > 3)** | **Reference negative** | **Total** |
|---|---|---|---|
| Predicted positive | TP = 15 | FP = 8 | 23 |
| Predicted negative | FN = 26 | TN = 51 | 77 |
| Total | 41 | 59 | 100 |

**Table S3.** Final selected radiomics features.

| **Feature** | **Selected in folds (of 5)** | **Mean coefficient** |
|---|---|---|
| adc_original_gldm_LargeDependenceHighGrayLevelEmphasis | 4 | 0.2242 |
| t2w_original_shape_Maximum2DDiameterRow | 3 | −0.0672 |
| t2w_original_glszm_LowGrayLevelZoneEmphasis | 3 | 0.0333 |

**Table S4.** Pairwise comparisons of model discrimination with Benjamini-Hochberg adjustment.

| **Comparison** | **Metric** | **Unadjusted P** | **Rank** | **BH-adjusted P** |
|---|---|---|---|---|
| Gland vs clinicopathologic | C-index | .0080 | 5 | .0160 |
| Gland vs lesion | C-index | .0100 | 6 | .0167 |
| Gland vs clinicopathologic | AUC at 24 months | <.0001 | 1 | <.0001 |
| Gland vs lesion | AUC at 24 months | .0040 | 4 | .0100 |
| Gland vs clinicopathologic | AUC at 36 months | <.0001 | 2 | <.0001 |
| Gland vs lesion | AUC at 36 months | .0020 | 3 | .0067 |
| Gland vs clinicopathologic | AUC at 48 months | .0140 | 7 | .0200 |
| Gland vs lesion | AUC at 48 months | .0500 | 10 | .0500 |
| Gland vs clinicopathologic | AUC at 60 months | .0160 | 8 | .0200 |
| Gland vs lesion | AUC at 60 months | .0340 | 9 | .0378 |

Note.—P values were derived from paired bootstrap resampling (1,000 iterations) and adjusted for multiple comparisons (n = 10) using the Benjamini-Hochberg procedure to control the false discovery rate at .05.

**Table S5.** Brier scores of the three models at 24, 36, 48 and 60 months.

| **Time point** | **Clinicopathologic** | **Lesion radiomics** | **Gland radiomics** |
|---|---|---|---|
| 24 months | 0.107 (0.073, 0.141) | 0.110 (0.076, 0.144) | 0.091 (0.067, 0.117) |
| 36 months | 0.110 (0.082, 0.137) | 0.110 (0.082, 0.140) | 0.089 (0.070, 0.109) |
| 48 months | 0.109 (0.087, 0.130) | 0.110 (0.086, 0.133) | 0.080 (0.065, 0.096) |
| 60 months | 0.099 (0.080, 0.117) | 0.099 (0.081, 0.119) | 0.077 (0.060, 0.095) |

Note.—Data are Brier score (95% CI); lower values indicate better performance.

## References

1. Raychaudhuri R, Lin DW, Montgomery RB. Prostate Cancer: A Review. JAMA. 2025;333(16):1433–1446. doi: 10.1001/jama.2025.0228.

2. Carlsson S, Benfante N, Alvim R, et al. Long-term outcomes of active surveillance for prostate cancer – the Memorial Sloan Kettering Cancer Center experience. J Urol. 2020;203(6):1122–1127. doi: 10.1097/JU.0000000000000713.

3. Hamdy FC, Donovan JL, Lane JA, et al. Fifteen-Year Outcomes after Monitoring, Surgery, or Radiotherapy for Prostate Cancer. N Engl J Med. Massachusetts Medical Society; 2023;388(17):1547–1558. doi: 10.1056/NEJMoa2214122.

4. Bokhorst LP, Lepistö I, Kakehi Y, et al. Complications after prostate biopsies in men on active surveillance and its effects on receiving further biopsies in the Prostate cancer Research International: Active Surveillance (PRIAS) study. BJU Int. 2016;118(3):366–371. doi: 10.1111/bju.13410.

5. Loeb S, Vellekoop A, Ahmed HU, et al. Systematic Review of Complications of Prostate Biopsy. Eur Urol. 2013;64(6):876–892. doi: 10.1016/j.eururo.2013.05.049.

6. Drost F-JH, Rannikko A, Valdagni R, et al. Can active surveillance really reduce the harms of overdiagnosing prostate cancer? A reflection of real life clinical practice in the PRIAS study. Transl Androl Urol. AME Publishing Company; 2018;7(1):9805–9105. doi: 10.21037/tau.2017.12.28.

7. Porten SP, Whitson JM, Cowan JE, et al. Changes in Prostate Cancer Grade on Serial Biopsy in Men Undergoing Active Surveillance. J Clin Oncol. Wolters Kluwer; 2011;29(20):2795– 2800. doi: 10.1200/JCO.2010.33.0134.

8. Lee C, Light A, Saveliev ES, Van Der Schaar M, Gnanapragasam VJ. Developing machine learning algorithms for dynamic estimation of progression during active surveillance for prostate cancer. Npj Digit Med. 2022;5(1):110. doi: 10.1038/s41746-022-00659-w.

9. Cooperberg MR, Zheng Y, Faino AV, et al. Tailoring Intensity of Active Surveillance for Low-Risk Prostate Cancer Based on Individualized Prediction of Risk Stability. JAMA Oncol. 2020;6(10):e203187. doi: 10.1001/jamaoncol.2020.3187.

10. Drost F-JH, Nieboer D, Morgan TM, et al. Predicting Biopsy Outcomes During Active Surveillance for Prostate Cancer: External Validation of the Canary Prostate Active Surveillance Study Risk Calculators in Five Large Active Surveillance Cohorts. Eur Urol. 2019;76(5):693–702. doi: 10.1016/j.eururo.2019.07.041.

11. Ankerst DP, Xia J, Thompson IM, et al. Precision Medicine in Active Surveillance for Prostate Cancer: Development of the Canary–Early Detection Research Network Active Surveillance Biopsy Risk Calculator. Eur Urol. 2015;68(6):1083–1088. doi: 10.1016/j.eururo.2015.03.023.

12. Tomer A, Nieboer D, Roobol MJ, et al. Personalised biopsy schedules based on risk of Gleason upgrading for patients with low-risk prostate cancer on active surveillance. BJU Int. 2021;127(1):96–107. doi: 10.1111/bju.15136.

13. Light A, Lophatananon A, Keates A, et al. Development and External Validation of the STRATified CANcer Surveillance (STRATCANS) Multivariable Model for Predicting Progression in Men with Newly Diagnosed Prostate Cancer Starting Active Surveillance. J Clin Med. Multidisciplinary Digital Publishing Institute; 2023;12(1):216. doi: 10.3390/jcm12010216.

14. Giganti F, Stabile A, Stavrinides V, et al. Natural history of prostate cancer on active surveillance: stratification by MRI using the PRECISE recommendations in a UK cohort. Eur Radiol. 2021;31(3):1644–1655. doi: 10.1007/s00330-020-07256-z.

15. Caglic I, Sushentsev N, Gnanapragasam VJ, et al. MRI-derived PRECISE scores for predicting pathologically-confirmed radiological progression in prostate cancer patients on active surveillance. Eur Radiol. 2021;31(5):2696–2705. doi: 10.1007/s00330-020-07336-0.

16. Midya A, Hiremath A, Huber J, et al. Delta radiomic patterns on serial bi-parametric MRI are associated with pathologic upgrading in prostate cancer patients on active surveillance: preliminary findings. Front Oncol. Frontiers; 2023;13. doi: 10.3389/fonc.2023.1166047.

17. Sushentsev N, Rundo L, Abrego L, et al. Time series radiomics for the prediction of prostate cancer progression in patients on active surveillance. Eur Radiol. 2023;33(6): 3792–3800. doi: 10.1007/s00330-023-09438-x.

18. Sushentsev N, Rundo L, Blyuss O, et al. Comparative performance of MRI-derived PRECISE scores and delta-radiomics models for the prediction of prostate cancer progression in patients on active surveillance. Eur Radiol. 2022;32(1):680–689. doi: 10.1007/s00330-021-08151-x.

19. Roest C, Kwee TC, Saha A, Fütterer JJ, Yakar D, Huisman H. AI-assisted biparametric MRI surveillance of prostate cancer: feasibility study. Eur Radiol. 2023;33(1):89–96. doi: 10.1007/s00330-022-09032-7.

20. Wang Y, Lou B. Predicting Prostate Cancer Progression During Active Surveillance Using Longitudinal bpMRI Scans and A Multi-scale Foundation Model. MIDL. 2025;

21. Wallaengen V, Zacharaki EI, Alhusseini M, et al. mpMRI-Based Risk Estimation to Optimize Prostate Cancer Patient Selection for Active Surveillance. Cancers. Multidisciplinary Digital Publishing Institute; 2026;18(5):842. doi: 10.3390/cancers18050842.

22. Sushentsev N, Rundo L, Blyuss O, Gnanapragasam VJ, Sala E, Barrett T. MRI-derived radiomics model for baseline prediction of prostate cancer progression on active surveillance. Sci Rep. Nature Publishing Group; 2021;11(1):12917. doi: 10.1038/s41598-021-92341-6.

23. Li CX, Bhattacharya I, Vesal S, et al. ProstAtlasDiff: Prostate cancer detection on MRI using Diffusion Probabilistic Models guided by population spatial cancer atlases. Med Image Anal. 2025;101:103486. doi: 10.1016/j.media.2025.103486.

24. Saha A, Bosma JS, Twilt JJ, et al. Artificial intelligence and radiologists in prostate cancer detection on MRI (PI-CAI): an international, paired, non-inferiority, confirmatory study. Lancet Oncol. Elsevier; 2024;25(7):879–887. doi: 10.1016/S1470-2045(24)00220-1.

25. Isensee F, Jaeger PF, Kohl SAA, Petersen J, Maier-Hein KH. nnU-Net: a self-configuring method for deep learning-based biomedical image segmentation. Nat Methods. Nature Publishing Group; 2021;18(2):203–211. doi: 10.1038/s41592-020-01008-z.

26. Chen T, Guestrin C. XGBoost: A Scalable Tree Boosting System. Proc 22nd ACM SIGKDD Int Conf Knowl Discov Data Min. New York, NY, USA: Association for Computing Machinery; 2016. p. 785–794. doi: 10.1145/2939672.2939785.

27. Lundberg SM, Lee S-I. A unified approach to interpreting model predictions. Proc 31st Int Conf Neural Inf Process Syst. Red Hook, NY, USA: Curran Associates Inc.; 2017. p. 4768–4777. https://dl.acm.org/doi/10.5555/3295222.3295230. Accessed June 11, 2026.

28. Li CX, Bhattacharya I, Vesal S, et al. Improving Automated Prostate Cancer Detection and Classification Accuracy with Multi-scale Cancer Information. In: Cao X, Xu X, Rekik I, Cui Z, Ouyang X, editors. Mach Learn Med Imaging. Cham: Springer Nature Switzerland; 2024. p. 341–350. doi: 10.1007/978-3-031-45673-2_34.

29. Rodrigues NM, Almeida JG de, Verde ASC, et al. Analysis of domain shift in whole prostate gland, zonal and lesions segmentation and detection, using multicentric retrospective data. Comput Biol Med. 2024;171:108216. doi: 10.1016/j.compbiomed.2024.108216.

30. Fassia M-K, Balasubramanian A, Woo S, et al. Deep Learning Prostate MRI Segmentation Accuracy and Robustness: A Systematic Review. Radiol Artif Intell. 2024;6(4):e230138. doi: 10.1148/ryai.230138.

## References

1. Saha A, Bosma JS, Twilt JJ, et al. Artificial intelligence and radiologists in prostate cancer detection on MRI (PI-CAI): an international, paired, non-inferiority, confirmatory study. Lancet Oncol. Elsevier; 2024;25(7):879–887. doi: 10.1016/S1470-2045(24)00220-1.

